# Improving risk assessment in forensic mental health: temporal validation and clinical refinement of the FoVOx risk tool

**DOI:** 10.64898/2026.01.20.26344471

**Authors:** Lenka Sivak, Jonas Forsman, Amir Sariaslan, Jari Tiihonen, Seena Fazel

## Abstract

**Background:** Forensic psychiatric services are expanding in many countries, and discharging patients from secure hospitals relies on accurate estimates of risk of adverse outcomes. Novel evidence-based tools for estimating one key risk, violent reoffending, have been developed in recent years. We aimed to externally validate one new tool, FoVOx, in forensic psychiatric patients sentenced to treatment, and to develop an updated model (FoVOx2), incorporating additional clinical predictors.

**Methods:** Using Swedish national registers, we conducted a temporal external validation of FoVOx by examining 767 patients discharged between 2014 and 2023. For the FoVOx2 cohort, 906 patients discharged between 2008 and 2023 were followed up, and additional predictors tested. The outcome was violent reconviction within 12 or 24 months. Model performance was evaluated using Harrell’s C-index, time-dependent AUCs, calibration, and classification metrics at predefined thresholds.

**Results:** In temporal validation, FoVOx showed moderate discrimination (AUCs 0.69 and 0.71; C-index = 0.69) and acceptable overall accuracy (Brier <0.11). Calibration was generally good, with mild overestimation at the highest predicted risks (>20%) at 12 months and slight underprediction at 24 months. The updated FoVOx2 model newly incorporated clozapine treatment and additional diagnostic categories. It was associated with improved performance (AUCs 0.77; optimism-corrected C-index = 0.72; Brier 0.06 and 0.09) and achieved good calibration (intercept ≈ 0; slopes 1.03 and 1.05).

**Conclusions:** Updating risk assessment tools with additional clinical factors can lead to incremental improvement in model performance. Implementing tools should consider clinical utility and impact as next steps.

## Introduction

Preventing violent recidivism (or repeat violent offending) after discharge from forensic psychiatric care is a key priority for patients and their carers, clinical services, and wider society. Rates of violent reoffending range from 3000 to 4500 offences per 100,000 person-years, which have been reported in national cohort studies [1–3]. Decisions about discharge from forensic services are based on many clinical and legal considerations, with a key one related to reoffending risks [4]. These decisions carry significant implications for resource allocation for community mental health services, and more widely for public health and safety, making accurate and transparent risk assessments an integral part of high-quality risk management and safe services [1, 5–12].

Traditional approaches relying on unstructured clinical judgment have shown limited reliability and predictive validity [13, 14]. In contrast, structured methods - professional judgment (SPJ) instruments and probabilistic (actuarial) models - provide more accurate and transparent risk estimates [15–17]. In particular actuarial tools, developed using empirically derived risk markers, offer standardized, time-efficient and consistent assessments with quantifiable risk estimates [18, 19]. They are especially useful in forensic psychiatry, where clear risk communication across judicial, social, and healthcare systems is necessary [20].

FoVOx was developed using Swedish data as a probabilistic risk tool for predicting violent reoffending following discharge from forensic psychiatric care [5]. The predictors that form the tools are routinely collected sociodemographic and clinical data, and intended to complement, rather than replace, structured clinical evaluations by providing a baseline level of risk [21]. The internally validated FoVOx model demonstrated good discriminatory accuracy (AUC = 0.77) in a nationwide cohort and was translated into a 12-item risk tool that is easy to implement and does not require specific training to use for clinicians. This made it well-suited for clinical environments where time and resources are limited [5, 22–24].

However, the original cohort is now more than a decade old, and included a heterogeneous sample of people legally sentenced to forensic psychiatric care and those treated by forensic psychiatric services in prison. The former population is mandated to undergo regular psychiatric monitoring and faces legally-sanctioned restrictions on discharge, which will alter reoffending risk [25, 26] – and is generalizable to patients in forensic psychiatric hospitals in most high-income countries (where people are sent to forensic hospitals under various legal sanctions, such as the Mental Health Act in England and Wales).

Therefore, this study had two aims: First, to conduct a temporal external validation of FoVOx among sentenced forensic psychiatric patients discharged between 2014 and 2023. Second, to update and internally validate the model (FoVOx2) using a more recent cohort of sentenced patients discharged between 2008 and 2023.

## Material and methods

### Data sources and linkage

Registers were cross-linked using Sweden’s unique 12-digit personal identity number, issued by the Swedish Tax Agency and used reliably across administrative and health registers [27]. The following databases were used:

- National Board of Forensic Medicine’s records: An internal database, covering all forensic psychiatric evaluations conducted in Sweden, available since 2009 [28].
- National Forensic Psychiatric Register (NFPR): A national quality registry, established in 2008, that monitors both inpatient and outpatient forensic psychiatric care, including diagnoses, treatments, and medication. In 2023, coverage included 96% of units and 84% of sentenced patients [26].
- National Patient Register (NPR): covering all inpatient admissions nationwide since 1987 [29].
- Longitudinal Integration Database for Health Insurance and Labour Market Studies (LISA): A comprehensive database containing annual socioeconomic information (e.g., age, sex, employment, education, income, benefits) since 1990.
- National Council for Crime Prevention’s Crime Register: Official data for convictions, including offence type, date of offence, and sentencing details [30].
- Cause of Death Register: A complete national register of all deaths since 1952.

### Study populations

The temporal validation cohort included individuals who underwent forensic psychiatric evaluation according to the National Board of Forensic Medicine’s records (since January 2009), were sentenced to forensic psychiatric care and later discharged between January 2014 and December 2023 (n = 767) according to NFPR. NFPR records both inpatient and outpatient compulsory care; hence “discharge” in this study denotes total discharge from forensic services (i.e., after outpatient part of their forensic care has also ended). This measure was chosen for both practical and clinical reasons: many patients transition repeatedly between inpatient and outpatient settings, and both forms of care constitute compulsory treatment with a high level of supervision, oversight, and control [25, 26]. For individuals with multiple treatment periods, one episode was randomly selected to avoid bias due to repeated observations. The updated cohort to develop a new model comprised sentenced patients totally discharged between November 2008 and December 2023 (n = 906). This larger cohort partially overlapped the validation sample but extended the timeframe to include earlier discharges and augment statistical power.

### Outcome

The primary outcome was conviction for a violent crime committed after total discharge within 12 and 24 months. Violent crimes included homicide, assault, robbery, arson, sexual offences, and unlawful threats or harassment [31]. The date of the offence was used as the event time to minimise bias from judicial processing delays.

### Predictors

Sociodemographic data, including age, sex, and employment status before admission, were obtained from LISA. Information on previous violent and serious violent crimes was extracted from the Crime Register. Serious violent crime was defined as homicide or manslaughter, aggravated assault, aggravated robbery, aggravated arson, rape, sexual coercion, or sexual exploitation. The NPR was used to identify the number of previous psychiatric inpatient treatments. The NFPR provided data on psychiatric and substance use diagnoses, psychotropic medication, and treatment duration. Diagnoses were classified to main categories using ICD-10: schizophrenia-spectrum disorders (F20–F29), bipolar disorder (F30–F31), unipolar depression (F32–F34.1), anxiety and stress-related disorders (F41, F43–F45, F48), antisocial personality disorder (F60.2), other personality disorders (F60-F62 excl. F60.2, F69), ADHD (F90), developmental disorders (F70-F79, F84), alcohol use disorder (F10) and drug use disorder (F11-F19). In the temporal validation, the original FoVOx model structure was maintained, in which a single primary diagnosis was used per patient. The new FoVOx2 model allowed for overlapping diagnoses to better reflect clinical practice where psychiatric comorbidities are common, and the assignment of a “primary” diagnosis can be inconsistent [32, 33]. In addition, for the updated model, we dropped two predictors: employment before admission and treatment duration shorter than 1 year as they were not statistically significant in preliminary analyses. The variable for number of previous inpatient episodes was simplified to a binary indicator (yes/no) to improve clinical feasibility and ease of implementation [23, 24].

### Model development and validation

For the temporal validation, the original FoVOx coefficients were applied to compute individual linear predictors and predicted risks of violent reoffending at 12 and 24 months after total discharge from compulsory forensic psychiatric care [5]. For model validation, missing data were handled using multiple imputation by chained equations (MICE) [34]. Data were censored for outcome, mortality, emigration or end of follow-up.

For the updated FoVOx2 model, the original FoVOx analytic protocol for predictor selection was used, employing a two-stage Cox proportional hazards approach. In the first stage, one set of predictors – sex, age at discharge, previous violent crime, schizophrenia-spectrum disorder, alcohol use disorder, and other substance use disorder – were included based on prior evidence and clinical relevance [3, 32, 35–37]. In the second stage, additional predictors were entered by stepwise selection until no variables remained with a p-value greater than 0.1 in multivariable models (aligning with original model development).

### Further statistical analyses

Discrimination was evaluated using two complementary metrics. The Harrell’s C-index measured the model’s overall ability to rank individuals by risk across the entire follow-up period (global concordance), inherently accounting for censoring. In addition, time-dependent area under the receiver operating characteristics curves (AUCs) were calculated at 12 and 24 months to assess discrimination at specific time horizons, using inverse probability of censoring weighting (IPCW) to adjust for right-censoring. IPCW is a weighting approach in which each individual’s contribution is scaled by the inverse of the estimated probability of not being censored at a given time, thus mitigating the right-censoring bias [38]. Calibration was assessed through calibration slopes and calibration plots comparing observed and predicted risks [39]. Overall calibration accuracy was estimated using IPCW-adjusted Brier scores [40].

Classification performance was summarised by sensitivity, specificity, positive predictive value (PPV), and negative predictive value (NPV) at prespecified risk thresholds of 5% and 20%, in line with the original FoVOx reporting. The proportional hazards assumption was tested using scaled Schoenfeld residuals. Internal validation of the new models was performed using bootstrap resampling (500 samples) to estimate and correct for optimism in performance metrics (C-index, calibration slope, Dxy, and R²). [41] Adequacy of our sample size was ensured by maintaining approximately 10 events of reoffending per candidate predictor – a recommended guideline to reduce the risk of overfitting in prediction modelling [42, 43].

All analyses were conducted using R version 4.4.2. The date of offence was used as the event time, and incomplete follow-up was handled by censoring, consistent with standard survival analysis methods [44]. In addition to the main analyses, tables and figures presented in this article, additional material – including a psychosis-specific model – is provided in the supplementary material.

### Ethics approval

Ethical approval for this study was obtained from the Swedish Ethical Review Authority (case number 2023-04161-01). All data were pseudonymised prior to analysis. Inclusion to the NFPR was based on an informed consent obtained at the time of first registration.

## Results

### Cohort characteristics

The temporal validation cohort comprised 767 individuals sentenced to forensic psychiatric care and fully discharged between January 2014 and December 2023. During follow-up, 118 patients (15.4%) were convicted of a new violent offence. The mean age at discharge was 42 years, and 79% were male. More than half (54.2%) suffered from schizophrenia-spectrum disorder, and approximately half (44.9%) had some form of substance use disorder. Median treatment duration was 54 months. The FoVOx2 development cohort included 906 sentenced patients discharged between November 2008 and December 2023, of whom 172 (19.0%) reoffended violently. Baseline characteristics for both cohorts are summarised in Tables 1 and 2.

**Table 1.**
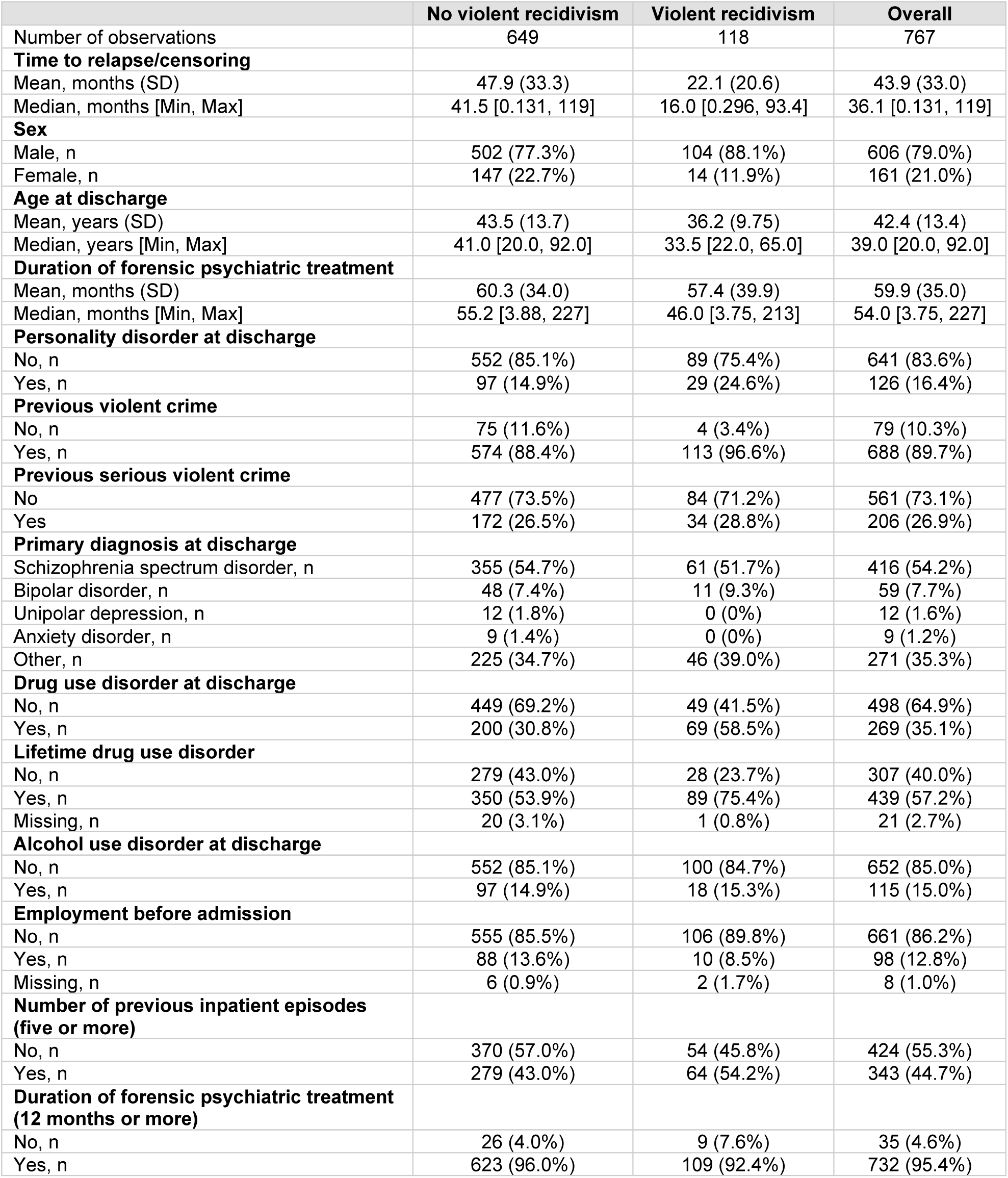
Temporal validation cohort. Individuals sentenced to forensic psychiatric treatment in Sweden, discharged between January 2014 and December 2023.

|  | No violent recidivism | Violent recidivism | Overall |
| --- | --- | --- | --- |
| Number of observations | 649 | 118 | 767 |
| **Time to relapse/censoring** |  |  |  |
| Mean, months (SD) | 47.9 (33.3) | 22.1 (20.6) | 43.9 (33.0) |
| Median, months [Min, Max] | 41.5 [0.131, 119] | 16.0 [0.296, 93.4] | 36.1 [0.131, 119] |
| **Sex** |  |  |  |
| Male, n | 502 (77.3%) | 104 (88.1%) | 606 (79.0%) |
| Female, n | 147 (22.7%) | 14 (11.9%) | 161 (21.0%) |
| **Age at discharge** |  |  |  |
| Mean, years (SD) | 43.5 (13.7) | 36.2 (9.75) | 42.4 (13.4) |
| Median, years [Min, Max] | 41.0 [20.0, 92.0] | 33.5 [22.0, 65.0] | 39.0 [20.0, 92.0] |
| **Duration of forensic psychiatric treatment** |  |  |  |
| Mean, months (SD) | 60.3 (34.0) | 57.4 (39.9) | 59.9 (35.0) |
| Median, months [Min, Max] | 55.2 [3.88, 227] | 46.0 [3.75, 213] | 54.0 [3.75, 227] |
| **Personality disorder at discharge** |  |  |  |
| No, n | 552 (85.1%) | 89 (75.4%) | 641 (83.6%) |
| Yes, n | 97 (14.9%) | 29 (24.6%) | 126 (16.4%) |
| **Previous violent crime** |  |  |  |
| No, n | 75 (11.6%) | 4 (3.4%) | 79 (10.3%) |
| Yes, n | 574 (88.4%) | 113 (96.6%) | 688 (89.7%) |
| **Previous serious violent crime** |  |  |  |
| No | 477 (73.5%) | 84 (71.2%) | 561 (73.1%) |
| Yes | 172 (26.5%) | 34 (28.8%) | 206 (26.9%) |
| **Primary diagnosis at discharge** |  |  |  |
| Schizophrenia spectrum disorder, n | 355 (54.7%) | 61 (51.7%) | 416 (54.2%) |
| Bipolar disorder, n | 48 (7.4%) | 11 (9.3%) | 59 (7.7%) |
| Unipolar depression, n | 12 (1.8%) | 0 (0%) | 12 (1.6%) |
| Anxiety disorder, n | 9 (1.4%) | 0 (0%) | 9 (1.2%) |
| Other, n | 225 (34.7%) | 46 (39.0%) | 271 (35.3%) |
| **Drug use disorder at discharge** |  |  |  |
| No, n | 449 (69.2%) | 49 (41.5%) | 498 (64.9%) |
| Yes, n | 200 (30.8%) | 69 (58.5%) | 269 (35.1%) |
| **Lifetime drug use disorder** |  |  |  |
| No, n | 279 (43.0%) | 28 (23.7%) | 307 (40.0%) |
| Yes, n | 350 (53.9%) | 89 (75.4%) | 439 (57.2%) |
| Missing, n | 20 (3.1%) | 1 (0.8%) | 21 (2.7%) |
| **Alcohol use disorder at discharge** |  |  |  |
| No, n | 552 (85.1%) | 100 (84.7%) | 652 (85.0%) |
| Yes, n | 97 (14.9%) | 18 (15.3%) | 115 (15.0%) |
| **Employment before admission** |  |  |  |
| No, n | 555 (85.5%) | 106 (89.8%) | 661 (86.2%) |
| Yes, n | 88 (13.6%) | 10 (8.5%) | 98 (12.8%) |
| Missing, n | 6 (0.9%) | 2 (1.7%) | 8 (1.0%) |
| **Number of previous inpatient episodes (five or more)** |  |  |  |
| No, n | 370 (57.0%) | 54 (45.8%) | 424 (55.3%) |
| Yes, n | 279 (43.0%) | 64 (54.2%) | 343 (44.7%) |
| **Duration of forensic psychiatric treatment (12 months or more)** |  |  |  |
| No, n | 26 (4.0%) | 9 (7.6%) | 35 (4.6%) |
| Yes, n | 623 (96.0%) | 109 (92.4%) | 732 (95.4%) |

**Table 2.** Updated cohort. Individuals sentenced to forensic psychiatric treatment in Sweden, discharged between November 2008 and December 2023.

|  | No violent recidivism | Violent recidivism | Overall |
| --- | --- | --- | --- |
| Number of observations | 734 | 172 | 906 |
| **Time to relapse/censoring** |  |  |  |
| Mean, months (SD) | 55.4 (40.5) | 28.5 (26.7) | 51.6 (40.1) |
| Median, months [Min, Max] | 47.6 [0.131, 173] | 20.3 [0.296, 132] | 39.8 [0.131, 173] |
| **Sex** |  |  |  |
| Male, n | 572 (77.9%) | 148 (86.0%) | 720 (79.5%) |
| Female, n | 162 (22.1%) | 24 (14.0%) | 186 (20.5%) |
| **Age at discharge** |  |  |  |
| Mean, years (SD) | 43.6 (13.7) | 37.4 (10.5) | 42.4 (13.4) |
| Median, years [Min, Max] | 41.0 [18.0, 92.0] | 35.0 [20.0, 68.0] | 40.0 [18.0, 92.0] |
| **Duration of forensic psychiatric treatment** |  |  |  |
| Mean, months (SD) | 56.1 (34.1) | 54.9 (39.1) | 55.9 (35.1) |
| Median, months [Min, Max] | 50.4 [2.17, 227] | 45.5 [3.75, 213] | 48.9 [2.17, 227] |
| **Previous violent crime** |  |  |  |
| No, n | 86 (11.7%) | 7 (4.1%) | 93 (10.3%) |
| Yes, n | 648 (88.3%) | 165 (95.9%) | 813 (89.7%) |
| **Previous serious violent crime** |  |  |  |
| No | 541 (73.7%) | 129 (75.0%) | 670 (74.0%) |
| Yes | 193 (26.3%) | 43 (25.0%) | 236 (26.0%) |
| **Schizophrenia spectrum disorder** |  |  |  |
| No, n | 212 (28.9%) | 48 (27.9%) | 260 (28.7%) |
| Yes, n | 522 (71.1%) | 124 (72.1%) | 646 (71.3%) |
| **Affective disorder** |  |  |  |
| No, n | 601 (81.9%) | 145 (84.3%) | 746 (82.3%) |
| Yes, n | 133 (18.1%) | 27 (15.7%) | 160 (17.7%) |
| **Developmental disorder (ID/ASD)** |  |  |  |
| No, n | 425 (57.9%) | 78 (45.3%) | 503 (55.5%) |
| Yes, n | 309 (42.1%) | 94 (54.7%) | 403 (44.5%) |
| **ADHD** |  |  |  |
| No, n | 652 (88.8%) | 143 (83.1%) | 795 (87.7%) |
| Yes, n | 82 (11.2%) | 29 (16.9%) | 111 (12.3%) |
| **Antisocial personality disorder** |  |  |  |
| No, n | 701 (95.5%) | 145 (84.3%) | 846 (93.4%) |
| Yes, n | 33 (4.5%) | 27 (15.7%) | 60 (6.6%) |
| **Personality disorder, other than antisocial** |  |  |  |
| No, n | 629 (85.7%) | 137 (79.7%) | 766 (84.5%) |
| Yes, n | 105 (14.3%) | 35 (20.3%) | 140 (15.5%) |
| **Drug use disorder** |  |  |  |
| No, n | 521 (71.0%) | 77 (44.8%) | 598 (66.0%) |
| Yes, n | 213 (29.0%) | 95 (55.2%) | 308 (34.0%) |
| **Alcohol use disorder** |  |  |  |
| No, n | 627 (85.4%) | 145 (84.3%) | 772 (85.2%) |
| Yes, n | 107 (14.6%) | 27 (15.7%) | 134 (14.8%) |
| **Previous psychiatric inpatient treatment** |  |  |  |
| No, n | 96 (13.1%) | 11 (6.4%) | 107 (11.8%) |
| Yes, n | 638 (86.9%) | 161 (93.6%) | 799 (88.2%) |
| **Long-acting injectables** |  |  |  |
| No, n | 484 (65.9%) | 122 (70.9%) | 606 (66.9%) |
| Yes, n | 250 (34.1%) | 50 (29.1%) | 300 (33.1%) |
| **Oral antipsychotics** |  |  |  |
| No, n | 521 (71.0%) | 122 (70.9%) | 643 (71.0%) |
| Yes, n | 213 (29.0%) | 50 (29.1%) | 263 (29.0%) |
| **Clozapine** |  |  |  |
| No, n | 645 (87.9%) | 166 (96.5%) | 811 (89.5%) |
| Yes, n | 89 (12.1%) | 6 (3.5%) | 95 (10.5%) |
| **Mood stabilizers** |  |  |  |
| No, n | 612 (83.4%) | 149 (86.6%) | 761 (84.0%) |
| Yes, n | 122 (16.6%) | 23 (13.4%) | 145 (16.0%) |
| **Antidepressants** |  |  |  |
| No, n | 515 (70.2%) | 138 (80.2%) | 653 (72.1%) |
| Yes, n | 219 (29.8%) | 34 (19.8%) | 253 (27.9%) |

### Temporal validation of FoVOx

In the sentenced forensic cohort, the original FoVOx model demonstrated moderately good discriminatory accuracy across the full follow-up period (C-index 0.69 [95% CI 0.64-0.73], as well as at fixed time horizons, with AUCs of 0.69 at 12 months and 0.71 at 24 months (Appendix Figure 1). Figure 1 and Appendix Figure 2 present the distribution of the linear predictor (LP) values by violent recidivism status. The distribution of predicted risks at fixed time points was concentrated toward the lower end, with most individuals below 10% and few exceeding 20% (Figure 2, Appendix Figure 3).

**Figure 1.**
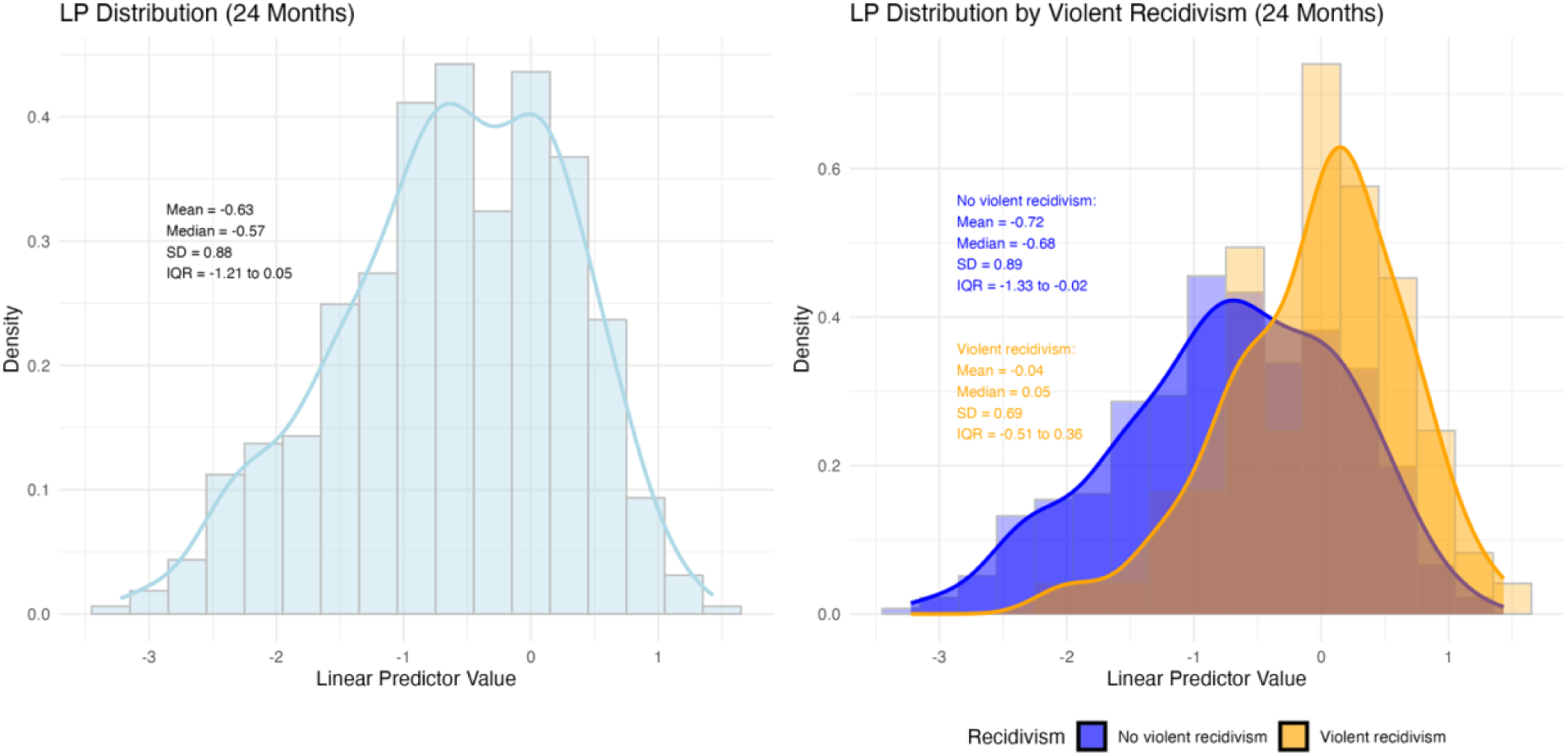
Distributions of the model’s linear predictor (LP) values for individuals with and without violent recidivism 24 months.

**Figure 2.**
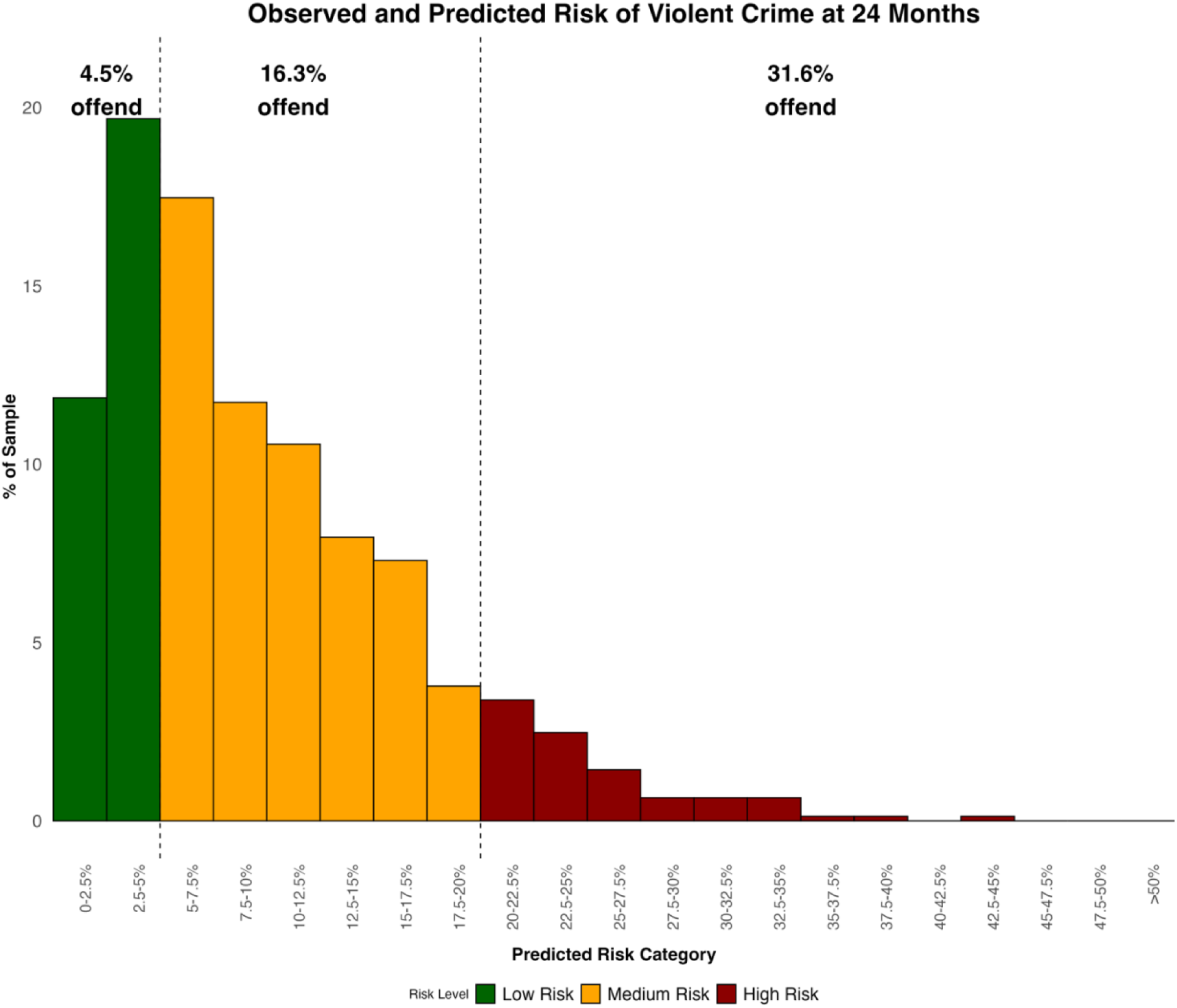
Observed and predicted risk of violent crime at 24 months, by risk category

**Figure 3.**
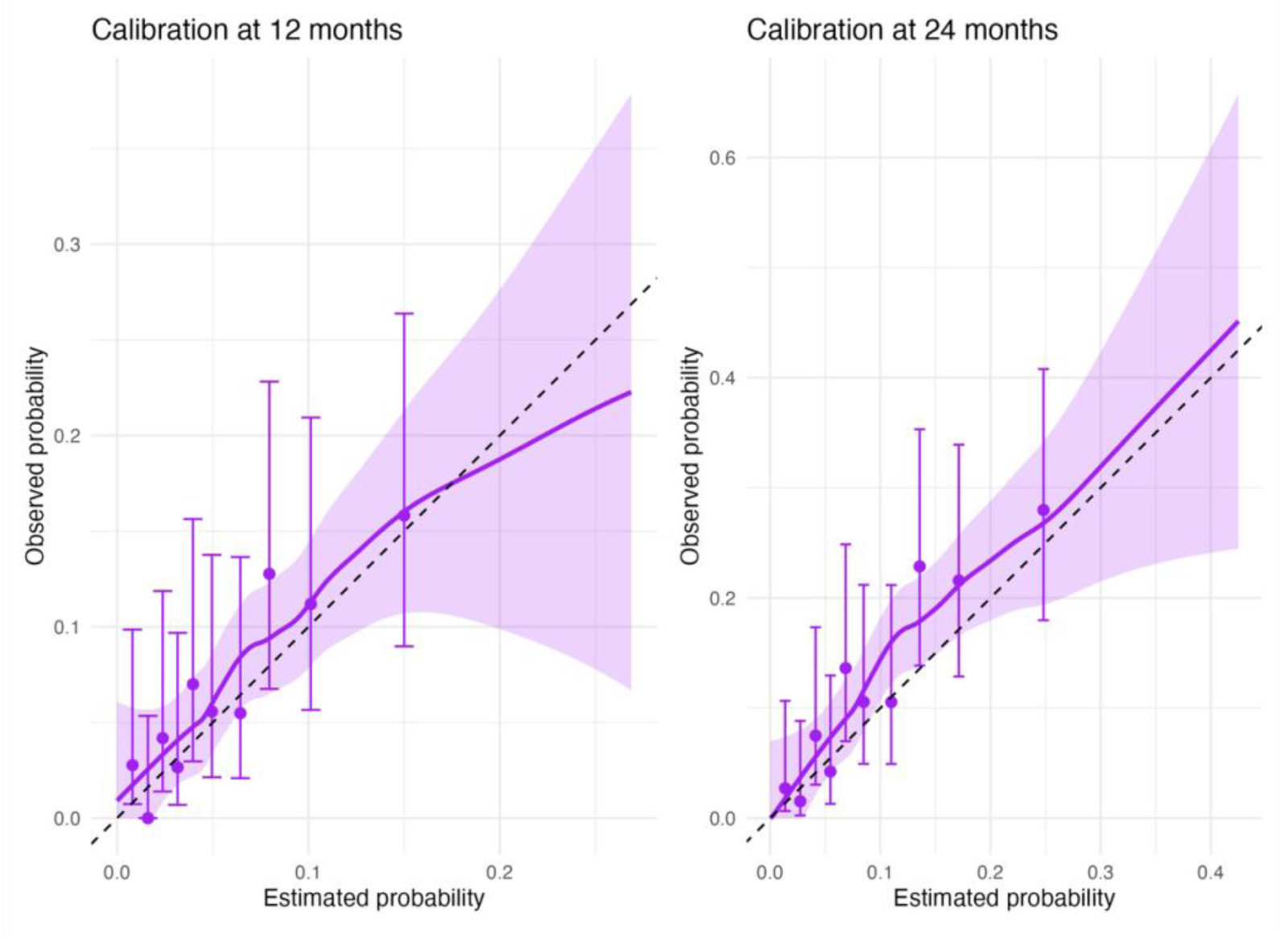
Calibration plots for FoVOx temporal validation.

Calibration was overall satisfactory. At 12 months, the model showed mild overprediction and somewhat extreme risk estimates (intercept -0.26; calibration slope 0.83). At 24 months, calibration was good, with an intercept of 0.20 and a slope of 0.95, indicating slight underprediction. Brier scores of 0.061 and 0.101 supported acceptable overall accuracy. At a 5% threshold, sensitivity/specificity were 0.70/0.57 (12 months) and 0.90/0.35 (24 months); at 20%, the trade-off reversed (0.02/0.99 and 0.22/0.93, respectively). Calibration plots (Figure 3) showed good alignment at lower predicted probabilities; at 12 months there was modest overestimation in the upper range, whereas at 24 months the model consistently underpredicted risk.

### FoVOx2: updated model and internal validation

In the new development cohort (n = 906, violent offences [events] = 172), the regression analyses identified additional independent predictors: antisocial personality disorder, other personality disorder, previous psychiatric inpatient treatment and clozapine treatment. Older age, female sex, absence of previous inpatient episodes and clozapine at discharge were associated with lower risk, whereas antisocial personality disorder, other types of personality disorders, drug use disorders, and previous violence markedly increased risk. Hazard ratios ranged from 0.3 (95% CI 0.1-0.6) for clozapine treatment to 3.9 (95% CI 1.8-8.3) for a history of violent offending (Table 3).

**Table 3.** Association between predictors and violent reoffending in the updated model (FoVOx2), derived using a two-stage Cox proportional hazards regression.

| Predictor | HR | 95% CI | p-value |
| --- | --- | --- | --- |
| Sex (female) | 0.61 | 0.39-0.95 | 0.029 |
| Age at discharge (per 1-yr older) | 0.97 | 0.95-0.98 | <0.001 |
| Previous violent crime | 3.90 | 1.82-8.35 | <0.001 |
| Schizophrenia spectrum disorder | 1.19 | 0.83-1.69 | 0.348 |
| Alcohol use disorder | 1.25 | 0.82-1.91 | 0.309 |
| Substance use disorder, other than alcohol | 2.34 | 1.70-3.22 | <0.001 |
| Antisocial personality disorder | 3.37 | 2.18-5.23 | <0.001 |
| Personality disorder, other than antisocial | 1.70 | 1.13-2.54 | 0.011 |
| Clozapine treatment | 0.28 | 0.12-0.63 | 0.002 |
| Absence of previous psychiatric inpatient care | 0.58 | 0.31-1.07 | 0.082 |

The model showed an apparent C-index of 0.75 (95% CI 0.71-0.78) and an optimism-corrected C-index of 0.72. Time-dependent AUCs were 0.77 (95% CI 0.70-0.83) at 12 months and 0.77 (95% CI 0.72-0-82) at 24 months (Figure 4). Calibration plots confirmed near-linear alignment up to approximately 50% predicted risk (Figure 5). Intercepts were -0.002 (95% CI -0.28-0.27) for 12 months and 0.03 (95% CI -0.20-0.25) at 24 months, suggesting no systematic miscalibration. Calibration slopes were 1.30 (95% CI 0.75-1.31) and 1.05 (95% CI 0.82-1.28), respectively. Brier scores (0.060 and 0.093) also indicated strong overall accuracy. Predicted risks were more dispersed for FoVOx2 than in the temporal validation set, with a heavier right-tail and a larger share of patients >0.20 risk probability (Figure 6, Appendix Figure 4). At the 20% thresholds, sensitivity/specificity were 0.23/0.96 at 12 months, and 0.46/0.86 at 24 months; corresponding metrics at the 5% threshold, as well as PPV and NPV, are provided in supplementary material (Appendix Table 3).

**Figure 4.**
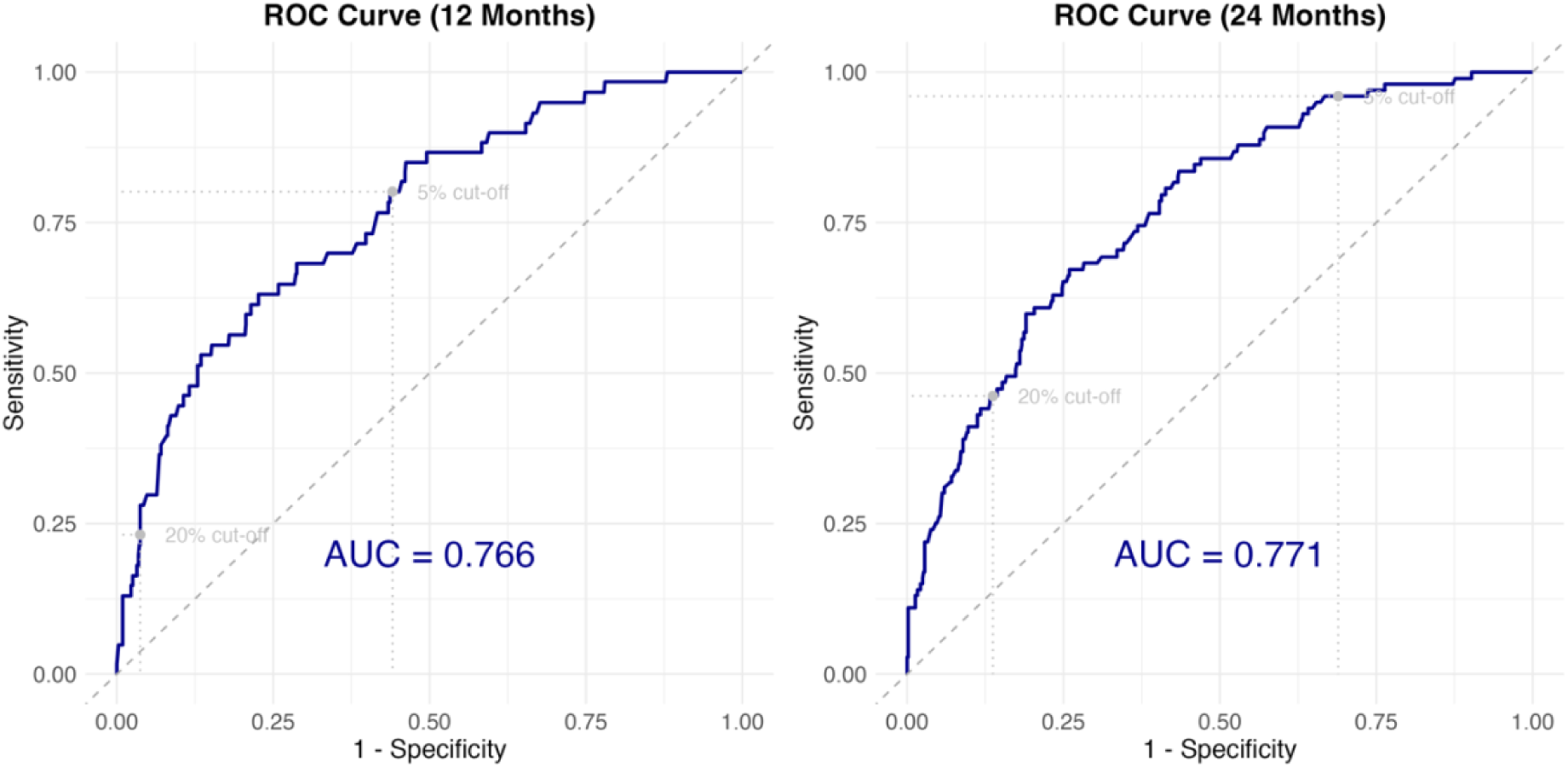
FoVOx2 model discrimination, presented as receiver operating characteristics (ROC) curves.

**Figure 5.**
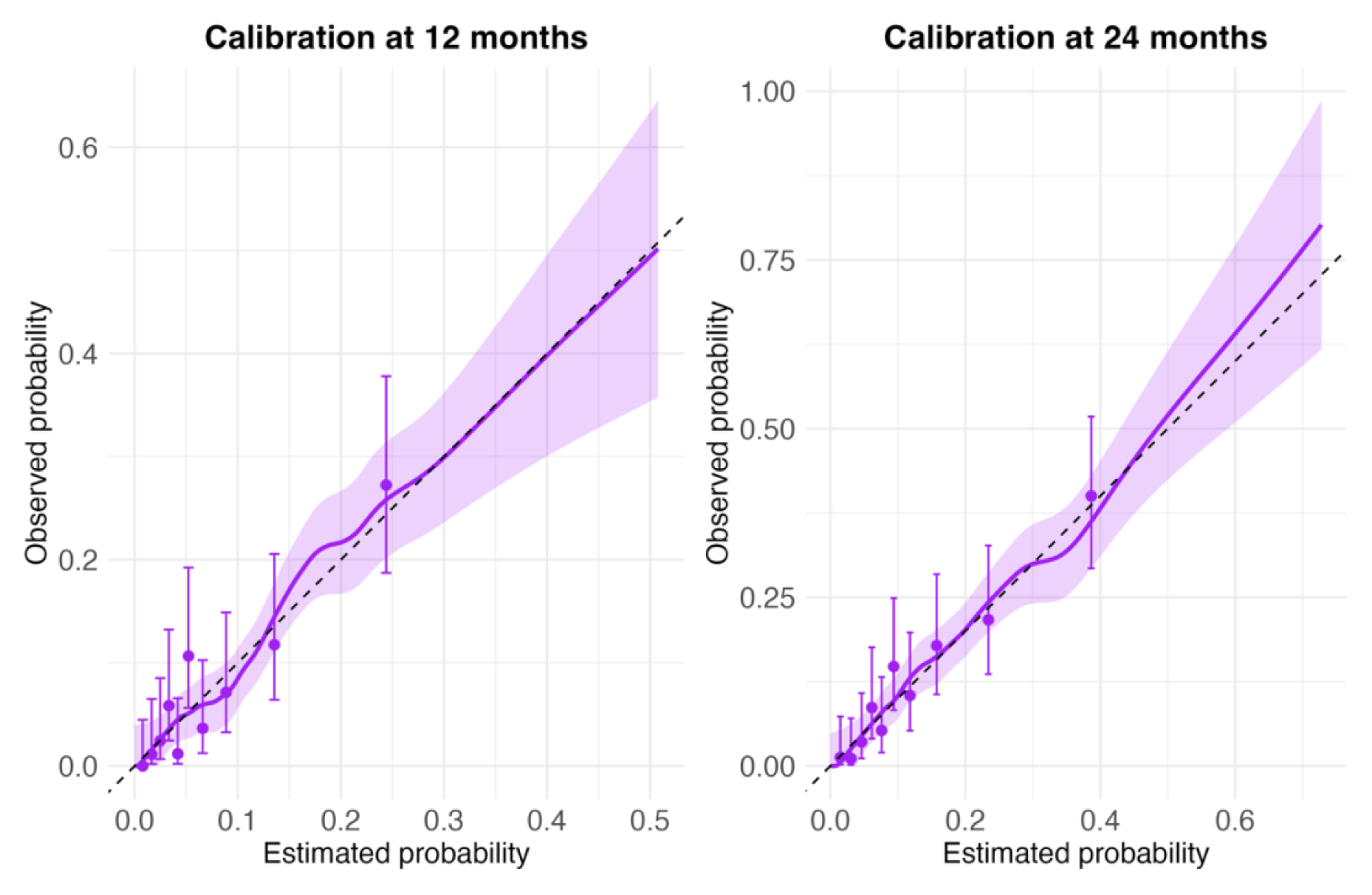
Calibration plots for FoVOx2 at 12 and 24 months.

**Figure 6.**
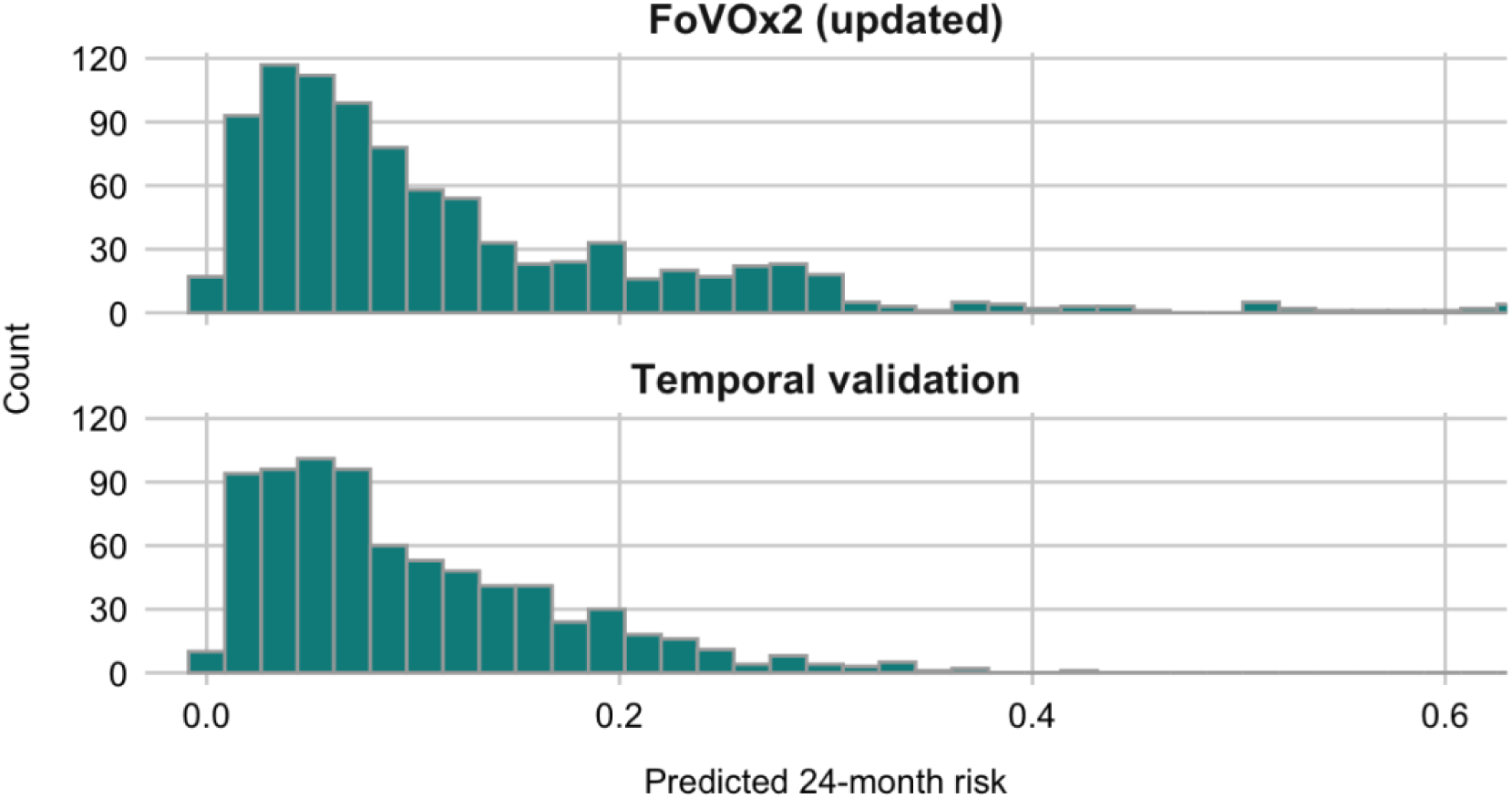
Predicted 24-month risk distributions: FoVOx2 (updated) vs. temporal validation.

## Discussion

Using nationwide Swedish register data, this study tested a new external validation of FoVOx – a novel scalable tool for predicting violent reoffending after discharge from forensic psychiatric care – and developed an updated version using additional clinical factors. In total, 906 patients were followed up for a mean of 52 months with 172 (19%) violent reoffences. The following principal findings emerged.

First, FoVOx demonstrated moderate to good performance in external validation, based on discrimination and calibration, when applied to a contemporary cohort of sentenced forensic patients. There was some shrinkage compared to the development sample (where AUCs were 0.77 at 12 and 24 months vs. AUCs of 0.69 and 0.71, respectively, in this external validation), which was not unexpected in a newer cohort. The findings indicate that FoVOx maintains predictive utility despite changes in clinical practice and patient mix over time. Reasons for this include the choice of baseline predictors, which was based on theory and empirical testing, and methodological aspects of the original study (which avoided overfitting, for example, and was sufficiently statistically powered).

Second, model updating was shown to be potentially useful. FoVOx2 did not include variables considered as difficult to assess reliably in routine practice, and added clinically relevant predictors, including clozapine treatment and two categories of personality disorder. These adjustments can additionally improve both the interpretability and feasibility of the tool. The inclusion of new treatment-related and diagnostic variables increased clinical relevance and better aligned the model with current forensic practice.

Third, incorporating these new predictors added incrementally to model performance. In the current sample, FoVOx2 achieved higher discrimination (AUCs 0.77) compared with the original model in the current validation (AUCs 0.69 and 0.71) and good overall calibration. Together, the study findings illustrate the value of updating actuarial tools and provide a model for testing the incremental value of new risk markers, ensuring utility as population and clinical practice evolve.

Another finding was that absence of previous psychiatric inpatient treatment was associated with lower risk of violent recidivism, which may reflect differences in underlying clinical trajectories within the forensic population. Individuals without prior inpatient care may include those whose offending occurred during an early or untreated phase of illness, with subsequent treatment leading to substantial risk reduction, whereas offending despite previous treatment may indicate a more persistent or complex risk profile, such as treatment resistance, antisocial traits or variable/non-adherence. For those with comorbid antisocial traits, further compulsory treatment may yield smaller reduction in recidivism risk.

### Clinical implications

With AUCs of 0.77, FoVOx2 demonstrated predictive performance comparable to or better than other validated violence risk models [15]. The updated models incorporated a medication variable (clozapine treatment), linking treatment status to reduced violence risk. Integrating medication data adds clinical depth and aligns with evidence that effective pharmacotherapy, particularly clozapine, can reduce violent behaviour in individuals with psychotic disorders [45, 46]. The updated instrument provides a transparent, data-driven, and reproducible alternative to structured professional judgement tools, which rely on subjective ratings and often show limited inter-rater reliability. The efficiency and feasibility of the new tool makes it well suited for routine clinical decision-making [23, 47].

A further consideration concerns communication of risk. Categorisation into “low”, “medium”, and “high” risk bands is practical when linked to clear risk levels but it can nevertheless obscure quantitative differences and potentially lead to arbitrary interpretation. Presenting predicted probabilities directly – as percentages – offers greater transparency and provide a common reference point for clinicians, courts, and decision-makers, helping contextualise risk without implying sharp thresholds or deterministic meanings. This approach aligns with current practice recommendations that actuarial tools complement, rather than replace, individualised clinical evaluation.

### Strengths and limitations

This study benefits from comprehensive national coverage, robust censoring methods, and large cohorts. Using offence dates rather than convictions minimised bias from judicial delays.

Limitations include reliance on administrative data, potential diagnostic misclassification, and partial temporal overlap between development and validation cohorts. Future external validations in independent and international forensic samples are needed to confirm generalisability. Feasibility studies can examine whether and how FoVOx2 can complement decision-making and can consider how to link risk ratings with management.

## Conclusion

In this nationwide temporal validation, the original FoVOx model maintained moderate discriminatory accuracy and calibration, with modest overprediction at the upper end of predicted risk at 12 months and slight underprediction at 24 months. The updated model, FoVOx2, demonstrated improvements in model performance over the original version of the tool. Our findings therefore extend the FoVOx framework to a more contemporary cohort of forensic patients and support its use as an empirically grounded and clinically interpretable tool for discharge planning and violence risk management in forensic mental health. Moving away from categorical labels toward reporting continuous probability estimates may further enhance interpretability and practical application, allowing risk estimates to augment clinical judgement rather than replacing it.

## Data Availability

The data supporting the findings of this study are not publicly available due to ethical and legal restrictions associated with sensitive information.

