## Appendix for "Improving risk assessment in forensic mental health: temporal validation and clinical refinement of the FoVOx risk tool"

**Appendix Table 1.** Coefficients of the original FoVOx model

| **Variable** | **Coefficient** |
|---|---|
| Sex (female) | -0.8407 |
| Age at discharge | -0.0299 |
| Previous violent crime | 1.1682 |
| Previous serious violent crime | -0.4480 |
| Primary diagnosis at discharge<br>- Schizophrenia spectrum<br>- Bipolar disorder<br>- Unipolar depression<br>- Anxiety disorders<br>- Other | <br>0<br>0.5994<br>0.2867<br>0.1142<br>0.304 |
| Drug use disorder at hospitalisation or discharge | -0.1188 |
| Alcohol use disorder at hospitalisation or discharge | 0.2288 |
| Personality disorder at discharge | 0.3052 |
| Employment before admission | -0.578 |
| Number of previous inpatient episodes (five or more) | -0.4676 |
| Lifetime drug use disorder | 0.7964 |
| Length of stay in forensic hospital (12 months or more) | -0.4576 |

LC (linear combination) = ∑ beta*value of risk factor

Risk of violent offending within 12 months = 1 – 0.9280^exp(LC)

Risk of violent offending within 24 months = 1 – 0.8762^exp(LC)

**Appendix Figure 1.** FoVOx temporal validation discrimination, presented as receiver operating characteristics (ROC) curves.

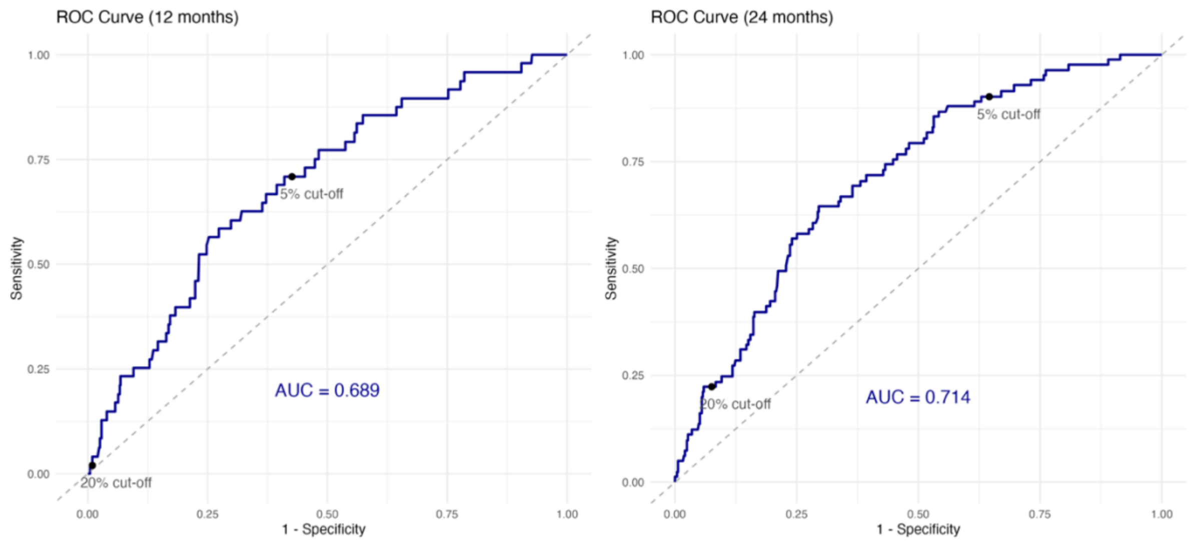


**Appendix Figure 2.** Distribution of the linear predictor (LP) values in the temporal validation sample at 12 months.

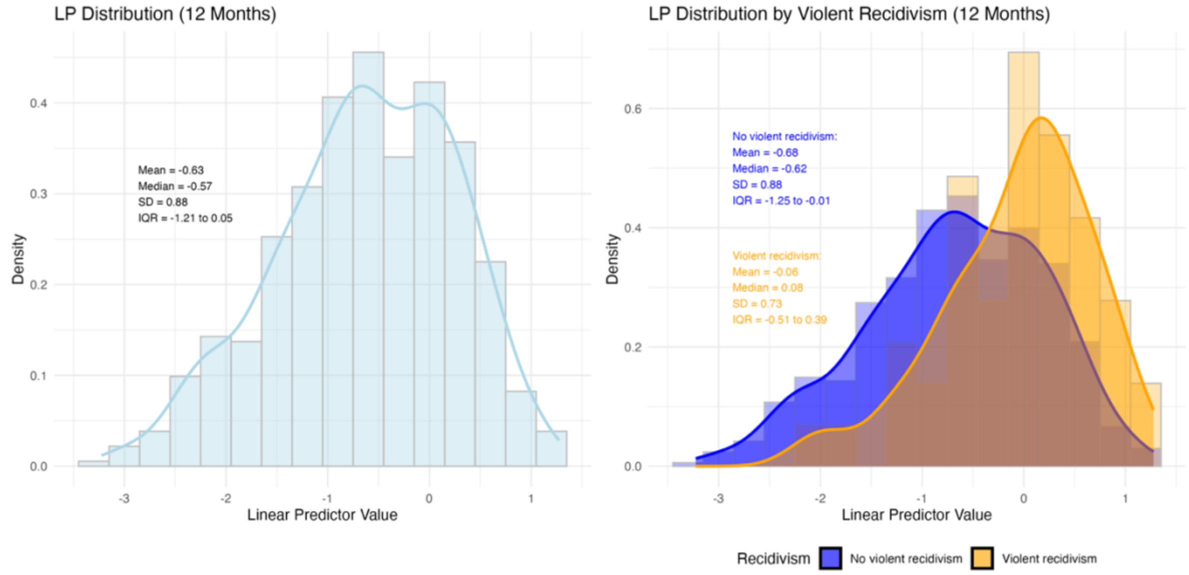

**Appendix Figure 3.** Observed and predicted risk of violent crime at 12 months in the temporal validation sample, by risk categorization.

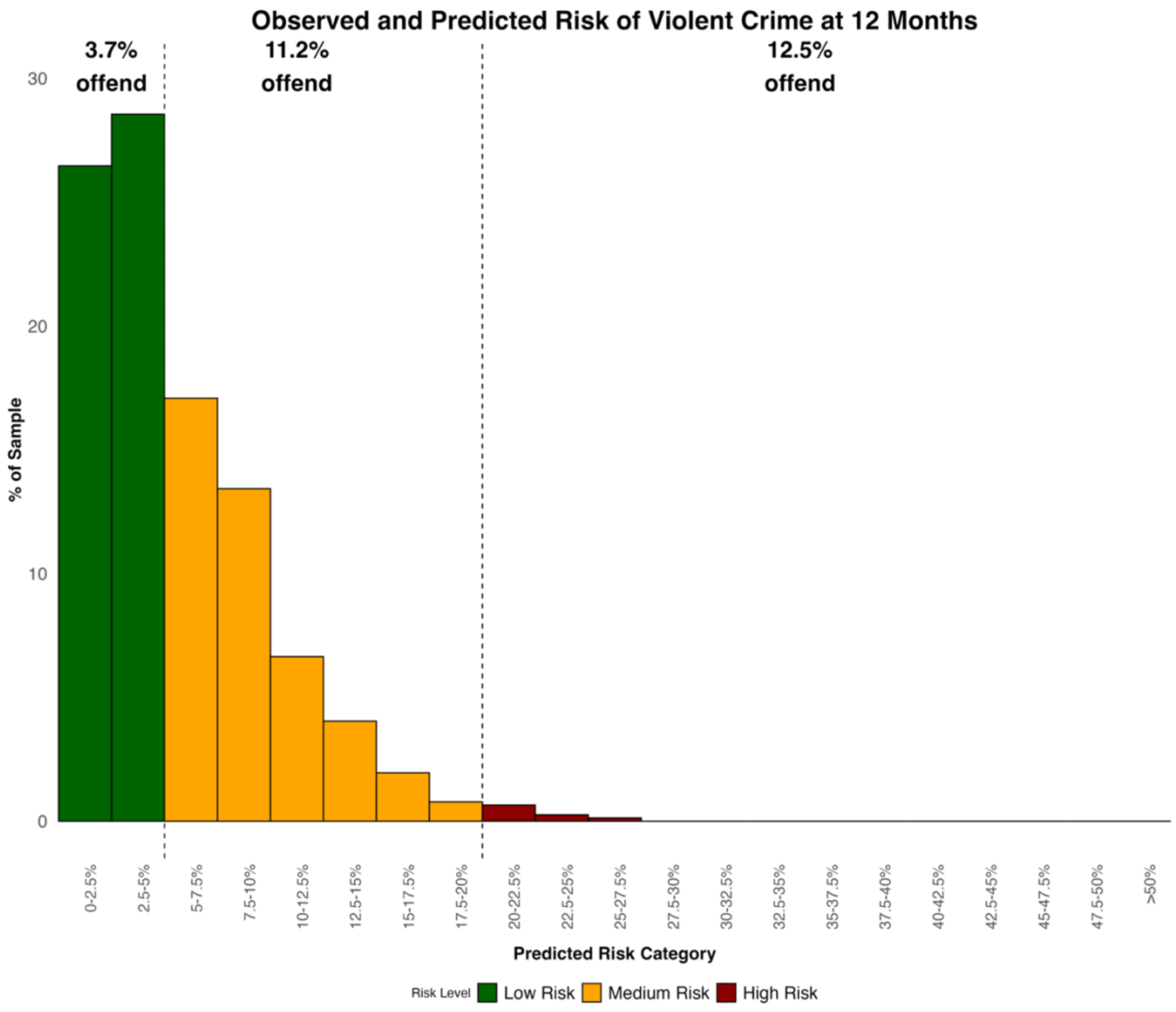

**Appendix Table 2.** Sensitivity, specificity, positive predictive value (PPV) and negative predictive value (NPV) for temporal validation at predefined thresholds at 12 and 24 months.

| **12 months** | | | | |
|---|---|---|---|---|
| Cut-off | Sensitivity | Specificity | PPV | NPV |
| 5% | 0.709 | 0.574 | 0.107 | 0.965 |
| 20% | 0.020 | 0.990 | 0.130 | 0.934 |
| **24 months** | | | | |
| Cut-off | Sensitivity | Specificity | PPV | NPV |
| 5% | 0.902 | 0.354 | 0.162 | 0.963 |
| 20% | 0.223 | 0.925 | 0.295 | 0.896 |

**Appendix Table 3.** Sensitivity, specificity, positive predictive value (PPV) and negative predictive value (NPV) for FoVOx2 at predefined thresholds: 12 and 24 months.

| **12 months** | | | | |
|---|---|---|---|---|
| Cut-off | Sensitivity | Specificity | PPV | NPV |
| 5% | 0.801 | 0.559 | 0.121 | 0.974 |
| 20% | 0.232 | 0.963 | 0.321 | 0.943 |
| **24 months** | | | | |
| Cut-off | Sensitivity | Specificity | PPV | NPV |
| 5% | 0.960 | 0.311 | 0.162 | 0.982 |
| 20% | 0.462 | 0.863 | 0.318 | 0.920 |

**Appendix Figure 4.** Predicted 12-month risk distributions: FoVOx2 (updated) vs. temporal validation.

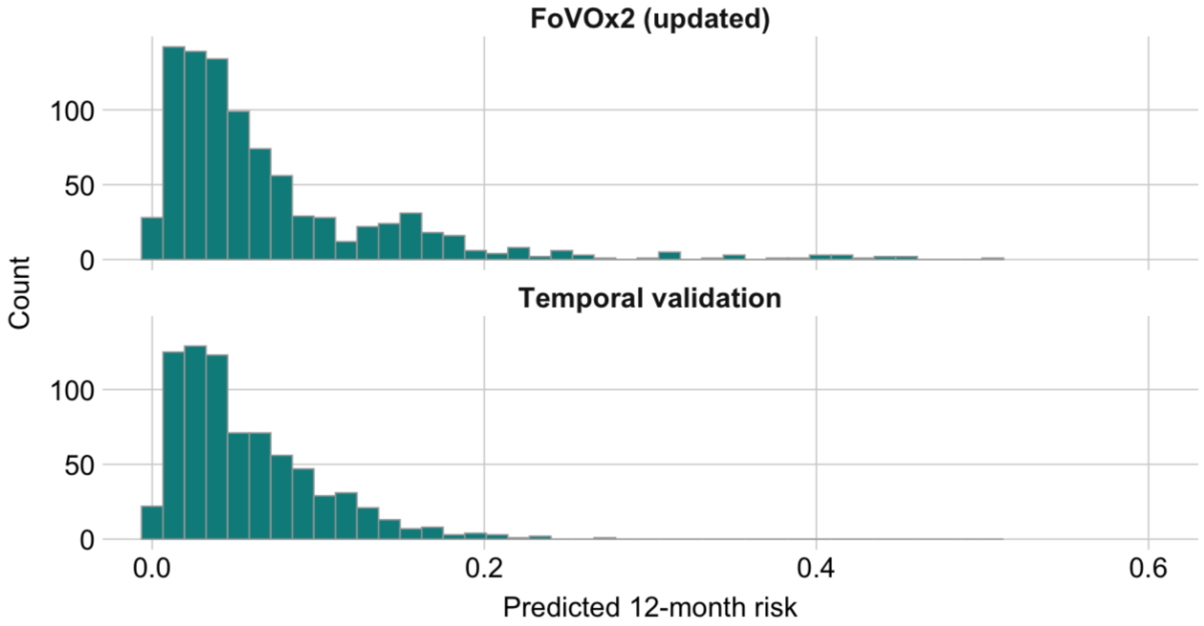


**Appendix Table 4.** FoVOx2 model coefficients

| **Predictor** | **Coefficient** |
|---|---|
| Sex (female) | -0.501525 |
| Age at discharge (per 1-yr older) | -0.033728 |
| Previous violent crime | 1.361418 |
| Schizophrenia spectrum disorder | 0.170431 |
| Alcohol use disorder | 0.220492 |
| Substance use disorder, other than alcohol | 0.849857 |
| Antisocial personality disorder | 1.216359 |
| Personality disorder, other than antisocial | 0.527684 |
| Clozapine treatment | -1.276121 |
| Absence of previous psychiatric inpatient care | -0.547761 |

LC (linear combination)=∑ beta*value of risk factor

Risk of violent offending within 12 months = 1 – 0.9898101^exp(LC)

Risk of violent offending within 24 months = 1 – 0.9813493^exp(LC)

**Appendix Text 1.** Psychosis-specific model FoVOx-P.

An additional aim of the current study was to develop a psychosis-specific model (FoVOx-P) applicable to jurisdictions where forensic psychiatric care is typically reserved to schizophrenia-spectrum disorders. Variables were selected using the Akaike Information Criterion (AIC) to identify the optimal combination of predictors. In the psychosis-restricted subsample (n = 646; events = 124), the FoVOx-P model selected following predictors: age, previous violent crime, substance use (alcohol and non-alcohol), antisocial personality disorder, clozapine treatment, absence of previous psychiatric inpatient treatment, and antidepressant medication. Younger age, previous violent offending, substance misuse, and antisocial traits were associated with elevated risk, while absence of previous inpatient care, clozapine and antidepressants were associated with reduced risk.

FoVOx-P achieved a C-index of 0.76 (SE = 0.02) and optimism-corrected 0.73. The optimism-corrected calibration slope was 0.82, suggesting moderate overfitting likely related to data-driven variable selection. AUCs were 0.77 (95%CI 0.70-0.85) at 12 months and 0.79 (95% CI 0.74-0.79) at 24 months. Brier scores were 0.061 and 0.093, and calibration remained satisfactory across all risk deciles.

**Appendix Table 5.** Association between predictors and violent crime in a psychosis-only sample model (FoVOx-P), derived using AIC-based Cox regression model.

| Predictor | HR | 95% CI | p-value |
|---|---|---|---|
| Age at discharge, (per 1-yr older) | 0.960 | 0.943-0.978 | <0.001 |
| Previous violent crime | 3.158 | 1.380-7.228 | 0.006 |
| Alcohol use disorder | 2.038 | 1.263-3.287 | 0.004 |
| Substance use disorder, other than alcohol | 2.382 | 1.627-3.487 | <0.001 |
| Antisocial personality disorder | 3.202 | 2.005-5.113 | <0.001 |
| Clozapine treatment | 0.244 | 0.099-0.602 | 0.002 |
| Antidepressants treatment | 0.627 | 0.378-1.040 | 0.071 |
| Absence of previous psychiatric inpatient care | 0.583 | 0.283-1.199 | 0.142 |

**Appendix Table 6.** FoVOx-P model coefficients.

| Predictor | Coefficient |
|---|---|
| Age at discharge (per 1-yr older) | -0.040312 |
| Previous violent crime | 1.149925 |
| Alcohol use disorder | 0.711914 |
| Substance use disorder, other than alcohol | 0.867888 |
| Antisocial personality disorder | 1.163771 |
| Clozapine treatment | -1.410993 |
| Antidepressants treatment | -0.466491 |
| Absence of previous psychiatric inpatient care | -0.540104 |

LC (linear combination)=∑ beta*value of risk factor

Risk of violent offending within 12 months = 1 – 0.985925^exp(LC)

Risk of violent offending within 24 months = 1 – 0.973313^exp(LC)

**Appendix Figure 6.** Calibration plots for FoVOx-P at 12 and 24 months.

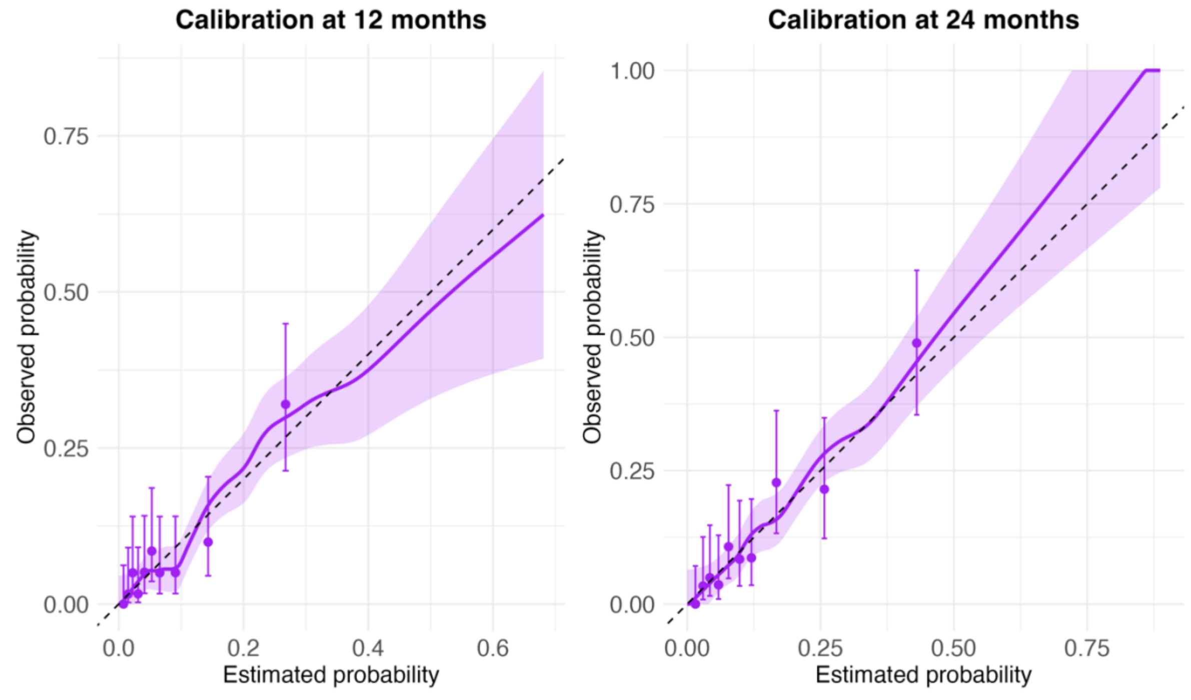
